# Longitudinal Bundibugyo Virus Glycoprotein Seroreactivity Following rVSVΔG-ZEBOV-GP Vaccination in Outbreak-Affected Populations of the Democratic Republic of the Congo

**DOI:** 10.64898/2026.06.22.26356273

**Authors:** Megan Halbrook, Sydney Merritt, Nicole A. Hoff, Patrick Mukadi, Jean Paul Kompany, Kamy Musene, Michael Beya, Handdy Kalengi, Merly Tambu, J. Daniel Kelly, Aquena H. Ball, Albert To, Christopher W. Woods, Bradly P. Nicholson, Micah T. McClain, Teri Wong, Lisa E. Hensley, Jason Kindrachuk, Axel T. Lehrer, Placide Mbala-Kingebeni, Anne W. Rimoin

## Abstract

**Background:** There are currently no vaccines approved for the prevention or treatment of Orthoebolavirus bundibugyoense (Bundibugyo virus; BDBV). The recombinant vesicular stomatitis virus–Zaire ebolavirus glycoprotein vaccine (rVSVΔG-ZEBOV-GP; ERVEBO) has been widely deployed during Ebola virus disease (EVD) outbreaks caused by Orthoebolavirus zairense (Ebola virus; EBOV). Given the lack of vaccines and medical countermeasures we evaluated development of antibodies to Bundibugyo glycoprotein (GP) following rVSVΔG-ZEBOV-GP vaccination in two EVD outbreak-affected populations in the Democratic Republic of the Congo (DRC).

**Methods:** Between 2018 and 2023, serum samples were collected from vaccine recipients in Mbandaka, Equateur Province (n=482 at baseline), and Beni, North Kivu Province (n=599 at baseline). Antibody reactivity was assessed using a multiplex pan-filovirus immunoassay. We evaluated longitudinal trends in BDBV GP seroreactivity across follow-up visits extending to approximately five years after vaccination.

**Findings:** We collected 2552 samples from 482 participants in Mbandaka and 3297 samples from 599 participants in Beni. BDBV GP responses diverged by location. Baseline BDBV GP seroreactivity differed between sites, with 3.3% of participants reactive in Mbandaka and 10.4% in Beni. In Mbandaka, BDBV GP titers remained unchanged through 6 months post-vaccination but increased markedly between 2.5 and 3.5 years (mean MFI 1,238 to 4,845; p<0.0001), accompanied by a rise in seroreactivity to 35.3%, followed by waning at later visits. In Beni, BDBV GP titers increased rapidly after vaccination, reaching peak seroreactivity at 21 days (52.3%) and 6 months (53.9%), with mean fold increases of 21.9 and 20.1 among baseline-naïve participants. Although antibody levels declined after 6 months, EBOV GP and BDBV GP titers remained above baseline, and significant increases in EBOV GP, BDBV GP, and SUDV GP titers were observed between 2.5 and 5 years.

**Interpretation:** We observed detectable BDBV GP seroreactivity in two geographically distinct populations in the DRC. However, there was no consistent evidence showing that rVSVΔG-ZEBOV-GP vaccination was associated with increased seroreactivity to BDBV GP across study populations. Baseline BDBV GP seroreactivity observed prior to vaccination warrants further investigation into the origins and epidemiological significance of pre-existing filovirus-reactive antibodies, including the possibility of prior exposure to related filoviruses, cross-reactive immune responses, high non-specific background reactivity unrelated to filoviruses, or other unrecognized sources of earlier antigenic stimulation. In the absence of licensed BDBV-specific vaccines, these data may inform preparedness planning, evaluation of existing countermeasures, and future vaccine development efforts during current and future BDBV outbreaks. It may further contribute to the study of natural history of filovirus immunity with a direct implication to understanding the possibility of cross-reactive and possible cross-protective responses in humans.

**Funding:** This project has been funded in whole or in part with Federal funds from the Food and Drug Administration (Grant No. 75F40119C10128) and the Gates Foundation (OPP1195609) and U.S. Defense Advanced Projects Agency (DARPA, N66001-09-C-2082 and HR0011-17-2-0069).

## INTRODUCTION

Prior to the Bundibugyo virus disease (BVD) outbreak declared in May 2026 in the Democratic Republic of the Congo (DRC) and Uganda, *Orthoebolavirus bundibugyoense* (Bundibugyo virus; BDBV) had been associated with two previously documented Bundibugyo disease outbreaks: the first in 2007 in Bundibugyo District, Uganda, a region bordering DRC, with a second in Isiro, DRC, in 2012^1,2^ (Figure 1). No licensed vaccines have been developed specifically for prevention of BVD, nor approved for use for other *Orthoebolavirus* species beyond *Orthoebolavirus zairens*e (EBOV). As public health authorities respond to this public health emergency of international concern, questions have emerged regarding whether existing Ebola vaccines, developed to primarily target the EBOV glycoprotein (GP), may generate immune responses that cross-protect against BDBV and other *Orthoebolaviruses*, and whether these responses could have relevance for future preparedness strategies.

**Figure 1:**
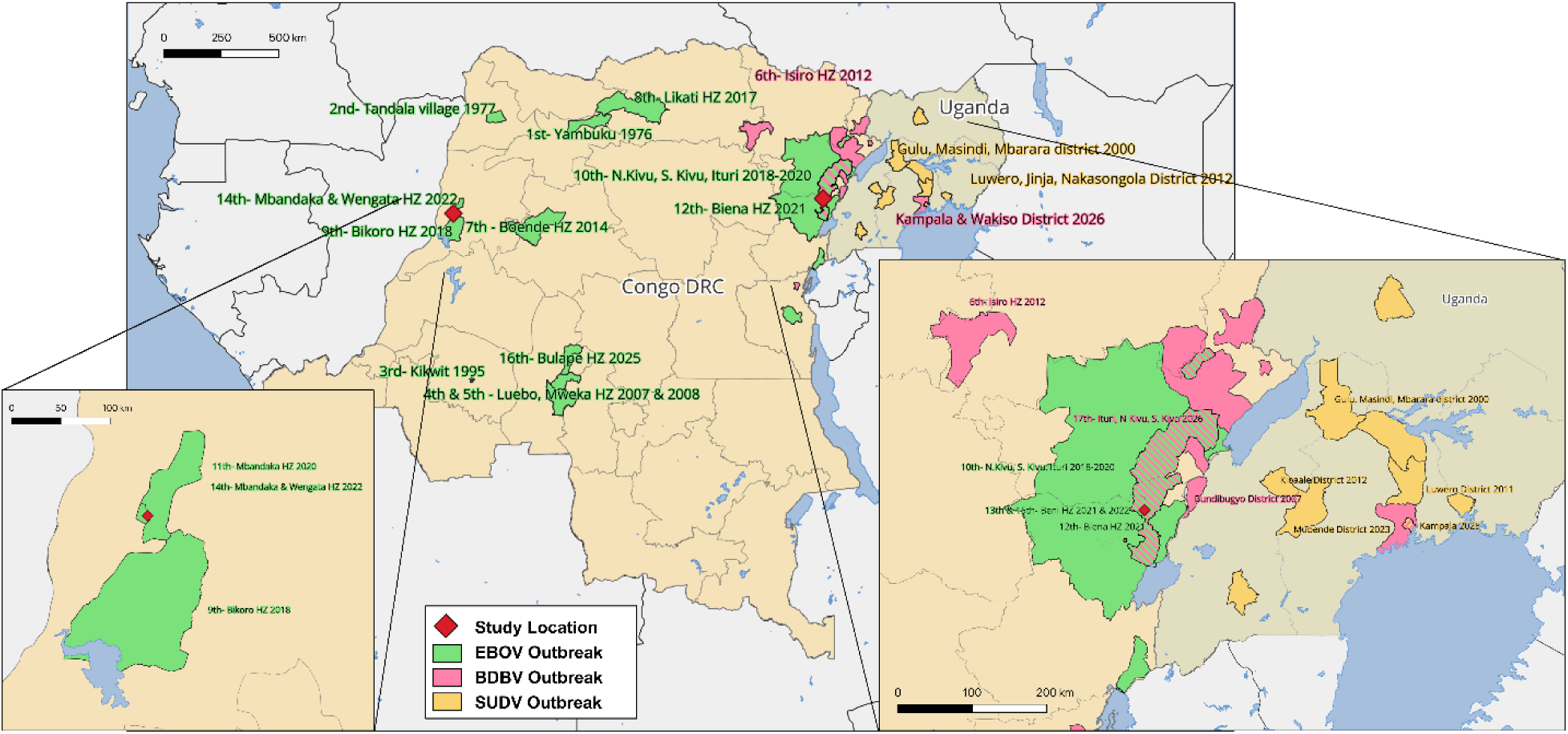
Map of past Ebola Outbreaks.

Ring-vaccination has become a central component of control strategies for Ebola Virus Disease (EVD) outbreaks. The recombinant vesicular stomatitis virus–EBOV glycoprotein vaccine (rVSVΔG-ZEBOV-GP; ERVEBO) demonstrated high efficacy and effectiveness against EVD in various outbreak settings and has been deployed during outbreak response activities beginning in 2015 in West Africa^3,4^. During the 2018–20 EVD outbreak in DRC, vaccination was found to reduce the risk of death in patients with EVD who were vaccinated before or after contact with an EVD patient, with effectiveness estimates of 84-98%^5–9^. During the 9th (Equateur province) and 10th (North Kivu, Ituri, South Kivu Provinces) EVD outbreaks beginning in 2018 in DRC, vaccination was implemented through ring-vaccination strategies targeting contacts, contacts of contacts, healthcare workers, and other frontline personnel. More than 303,000 individuals received vaccination during this response, generating one of the largest populations of ERVEBO vaccinated individuals living in regions with ongoing risk of filovirus emergence^10^.

Evidence regarding heterologous immune responses towards other Ebola species following rVSVΔG-ZEBOV-GP vaccination remains limited. A recent study of the J&J/Janssen Ebola virus vaccine (Ad26.ZEBOV) and 8-week MVA-BN-Filo booster administered in five African countries found that immune responses were specific to EBOV, despite the multivalent nature of the booster component^11^. Current available human data is sparse and largely derived from clinical trial populations evaluated over relatively short periods following vaccination, providing limited information on the effectiveness or durability of these responses following vaccine deployment during outbreaks. While a biologic basis for cross-reactive immune recognition may exist, the ability of current vaccines to offer cross-immunity remains unknown. Using an alternative multiplexed immunoassay approach, a recent study detected limited anti-BDBV seroreactivity at 28 days and 3 months post-immunization with rVSVΔG-ZEBOV-GP among a cohort of 72 individuals in Sierra Leone, Guinea, and Mali^12^. While there is considerable interest in vaccines that are broadly protective across the *Orthoebolavirus* genus, data addressing cross-reactivity in humans is limited and cross-protection in humans is unknown.

We established longitudinal cohorts in Mbandaka, Equateur Province and Beni, North Kivu Province, to evaluate immune responses following deployment of rVSVΔG-ZEBOV-GP with an Emergency Use Authorization (EUA) during response activities to two distinct EVD outbreaks in each province beginning in 2018^13–15^. Participants were enrolled during ring vaccination campaigns initiated during these outbreaks and followed for approximately five years.

## METHODS

### Study design and participants

Recruitment and enrollment of this study coincided with the rVSVΔG-ZEBOV-GP vaccination efforts undertaken in response to the DRC’s 9th and 10th EVD outbreaks. Beginning June 2018 in Mbandaka, Equateur, and in August, 2018 in Beni, North Kivu, individuals ≥1 year of age who received the rVSVΔG-ZEBOV-GP vaccine under the World Health Organization (WHO) ring vaccination protocol were eligible for enrollment^16^. These populations were defined as: confirmed EVD cases, contacts of cases or contacts-of-contacts, and healthcare or frontline workers in EVD affected areas. While pregnant women were included in the vaccination strategy, they were excluded from participation in this study. Recruitment and enrollment took place shortly after vaccination to limit disruptions to the vaccine deployment led by the Expanded Program for Immunization (EPI) of the Ministry of Health and WHO.

In Mbandaka, enrollment began on 07-Jun-2018 through 03-Jul-2018, and enrollment in Beni began on 15-Aug-2018 through 29-Aug-2018; all participants were enrolled on the day of vaccination with rVSVΔG-ZEBOV-GP. Both cohorts were followed over the course of five years, with seven collection periods in each province. Visit numbers are labeled by the estimated time from baseline, subject to logistic feasibility for field visits; exact follow up dates are presented in Table 1. In Beni, a study visit was made 1.5 years after vaccination and in Mbandaka a study visit was conducted 4.2 years after vaccination, coinciding with a disbursement of a booster dose of rVSVΔG-ZEBOV-GP vaccine which was given based on WHO booster recommendations.

**Table 1.** Study Visit Start Dates.

| **Visit** |  | **Mbandaka, Equateur** | **Beni, North Kivu** |
| --- | --- | --- | --- |
| Baseline |  | 2-Jun-18 | 15-Aug-18 |
| Follow Up Visit 1 | ~21 Days | 20-Jun-18 | 4-Sep-18 |
| Follow Up Visit 2 | ~6 Months | 5-Feb-19 | 22-Mar-19 |
| Follow Up Visit 3 | ~1.5 Years | - | 4-Jan-20 |
| Follow Up Visit 4 | ~2.5 Years | 8-Dec-20 | 14-Dec-20 |
| Follow Up Visit 5 | ~3.5 Years | 15-Mar-22 | 30-Apr-22 |
| Follow Up Visit 6 | ~4.2 Years | 22-Feb-23 | - |
| Follow Up Visit 7 | ~5 Years | 25-Aug-23 | 1-Dec-23 |

Over the five-year course of this study, our study regions experienced two additional EBOV outbreaks in Equateur (01-Jun-2020 to 18-Nov-2020 and 23-Apr-2022 to 4-Jul-2022) and three additional outbreaks in North Kivu (07-Feb-2021 to 3-May-2021, 08-Oct-2021 to 16-Dec-2021, 22-Aug-2022 to 27-Sep-2022). On two occasions in Mbandaka, study visits closely aligned with the start or end of an EBOV outbreak. Visit 4 (2.5 years post-vaccination) took place less than a month after DRC’s 11th outbreak.

In the weeks following data collection for Visit 5 (3.5 years post-vaccination), it was announced that an EBOV outbreak including five confirmed cases had been identified in Wangata and Mbandaka health zones, which overlapped with our study collection sites (Wangata, Mbandaka, and Bolenge). This was classified as the 14th Ebola disease outbreak reported in the DRC. Viral genome sequencing for both outbreaks suggested that they were due to new emergence events and not linked to previous outbreaks^17^.

At each study visit, participants completed an investigator-led survey, a short physical assessment, and provided a blood sample collected in red top serum tubes^18^. All blood samples were centrifuged, and heat inactivated for 1 hour at 56C for transport and storage; 1mL serum aliquots were stored at −20 °C in field laboratories and transferred to −80 °C storage at the National Institute for Biomedical Research (INRB) in Kinshasa.

### Laboratory analysis

A pan-filovirus multiplex immunoassay (MIA) was used for serological testing of all samples. This MIA included the following purified recombinant targets: EBOV GP, EBOV nucleoprotein (NP), EBOV viral protein 40 (VP40)^19,20^, BDBV GP, SUDV GP (Mayerlen et al. 2025), MARV GP and MARV VP40. All antigens were expressed using the drosophila expression system, produced and coupled to Magplex beads at the University of Hawaii. In addition to previously described antigens, gene sequences encoding amino acids 1–651 of BDBV Butalya strain (2007) glycoprotein (GP) (Genbank accession number YP_003815435) with a factor X cleavage site (aa IEGR), a short glycine linker (aa GGGS), followed by a c-terminal C-tag (aa EPEA), and gene sequences encoding amino acids 1-303 of MARV Musoke strain VP40 (Genbank accession number YP_001531155) further containing a C-terminal his-tag were inserted into pMT/BiP (Thermo Fisher Scientific, Waltham, MA). These vectors were co-transformed with the selectable marker plasmid pCoHygro into Drosophila S2 cells adapted to ExCell420 medium (Sigma-Aldrich, St. Louis, MO). Stable transformants were subsequently selected by the addition of 300 μg/mL hygromycin B (InvivoGen, San Diego, CA) to the medium. Expression of all GP proteins was induced by CuSO4 addition to the medium. The BDBV GP was purified using a CaptureSelect™C-tag column (Thermofisher, Waltham, MA) and the MARV VP40 was purified using a 2-step cation exchange using a HiTrap Capto Sp Imp Res column (GE Healthcare, Piscataway, NJ) followed by nickel ion-affinity chromatography with a HisTrap column (GE Healthcare, Piscataway, NJ). Bovine serum albumin (BSA)-coupled beads were used as an indicator of background reactivity. MIA methods have been previously described^21^; in brief, samples were diluted to 1:100 using 2 µL of serum and run in duplicate with an output of median fluorescent intensity (MFI) for each antigen target. Before analysis, detected anti-BSA background MFI was subtracted from the mean MFI for each filoviral antigen for each individual, with a limit of 0 MFI to avoid negative values. Seroreactivity thresholds specific to the DRC have been previously determined as the mean MFI plus three standard deviations of a filovirus Kinshasa cohort after BSA-adjustment (n = 379^19^). For all targets, binary seroreactivity at any time point was determined if the BSA-adjusted MFI value of the observation was greater than the Kinshasa-naive cut off as previously described^19^. The reactivity of EBOV GP for this assay has been compared to the Filovirus Animal Non-clinical Group (FANG) assay, which is considered a gold-standard ELISA, and a strong correlation in detected antibody reactivity was demonstrated in this comparison^22^.

### Statistical analysis

Statistical assessments of MFI value differences across timepoints was assessed using Welch’s t-test for summary statistics or ANOVA. Analyses were conducted in R (version 4.5.2, R Core Team, 2026). Maps were created using QGIS (QGIS Development Team, 2026).

### Ethical Considerations

All study activities were conducted under approval from appropriate institutional review boards: University of California, Los Angeles IRB No. IRB-16-001346 and IRB-20-001091, and Kinshasa School of Public Health IRB No. ESPCE0222017. Secondary analysis was conducted using a de-identified dataset with serologic results from all consenting individuals. Additionally, the study was approved by the Scientific Committee for Ebola Research during an outbreak at the INRB under the Ministry of Health. Prior to data or serum collection, participants signed or marked the approved informed consent form, and parents or guardians provided this consent on behalf of all child participants, while adolescents aged 7 to 17 provided assent as appropriate.

## RESULTS

During our five-year observation period from 2018-2023, we collected 2,552 serum samples from 482 participants in Mbandaka and 3,297 serum samples from 599 participants in Beni. Overall, participants in Mbandaka and Beni were primarily 19-55 years of age (>80%); 7.5% and 6.8% were 6-18 years old in Mbandaka and Beni, respectively, and no participants less than 6 years of age were enrolled (Table 2). Participants were enrolled from three health zones in the areas surrounding both Mbandaka and Beni. Participants were primarily male: 68.9% in Mbandaka and 63.9% in Beni. One third of the participants in each cohort were healthcare workers (33.2% and 34.9%).

**Table 2.** Participant Demographics.

|  | Mbandaka, Equateur |  | Beni, North Kivu |  |
| --- | --- | --- | --- | --- |
|  | N = 482 |  | N= 599 |  |
|  | N | % | N | % |
| **Age** |  |  |  |  |
| 0-5 years old | 0 | 0 | 0 | 0 |
| 6- 18 years old | 36 | 7.5 | 41 | 6.8 |
| 19-35 years old | 152 | 31.5 | 314 | 52.4 |
| 36-55 years old | 241 | 50.0 | 191 | 31.9 |
| 56-82 years old | 53 | 11.0 | 53 | 8.8 |
| **Health zone** |  |  |  |  |
| Bolenge | 100 | 20.7 |  |  |
| Mbandaka | 146 | 30.3 |  |  |
| Wangata | 236 | 49 |  |  |
| Beni |  |  | 234 | 39.1 |
| Mangina |  |  | 250 | 41.7 |
| Oicha |  |  | 99 | 16.5 |
| Other |  |  | 16 | 2.7 |
| **Sex** |  |  |  |  |
| Male | 332 | 68.9 | 383 | 63.9 |
| Female | 150 | 31.1 | 216 | 36.1 |
| **Occupational Exposures\*** |  |  |  |  |
| Healthcare Worker | 160 | 33.2 | 209 | 34.9 |
| Zoonotic Exposure | 11 | 2.3 | 9 | 1.5 |
| Other | 145 | 30.1 | 198 | 33.1 |
| Unknown | 166 | 34.4 | 183 | 30.6 |
| **Vaccination Criteria\*\*** |  |  |  |  |
| Close Contact of Ebola Patient | 76 | 15.8 | 237 | 39.6 |
| Healthcare Worker | 154 | 32.0 | 112 | 18.7 |
| Contact-of-Contact of Ebola Patient | 39 | 8.1 | 177 | 29.5 |
| Other | 213 | 44.2 | 73 | 12.2 |
| **Study Visit Participation** |  |  |  |  |
| Baseline | 482 | 100.0 | 599 | 100.0 |
| 21 Days | 467 | 96.9 | 541 | 90.3 |
| 6 Months | 417 | 86.5 | 501 | 83.6 |
| 1.5 Years |  |  | 467 | 78.0 |
| 2.5 Years | 355 | 73.7 | 373 | 62.3 |
| 3.5 Years | 329 | 68.3 | 391 | 65.3 |
| 4.2 Years | 198 | 41.1 |  |  |
| 5 Years | 304 | 63.1 | 425 | 71.0 |

\* Occupational exposure is derived from participant reported occupation. Participants reporting livestock farming or hunting as their primary occupation were categorised as having zoonotic occupational exposure.
\*\* Inclusion criteria for rVSVΔG-ZEBOV-GP ring vaccination was close contacts of a case, contact-of-contacts, healthcare workers, and in several situations other persons of risk. As persons could qualify for more than one criteria we sorted them into the category of highest risk- 23% of persons from Mbandaka and 46% from Beni. Our schema is: close contact > healthcare worker > contact of the contact > other.

Following vaccination with rVSVΔG-ZEBOV-GP, we observed a sustained increase in the mean anti-EBOV GP antibody MFI values over the 5-year period in both study locations^23^. Beyond that, there was little similarity in the trajectory of other filovirus antibody MFI values over time in these two locations.

At baseline collection in the **Mbandaka cohort,** 3.3% (n=16) participants were observed to be seroreactive to BDBV GP and 28.8% (n=139) participants were reactive to EBOV GP. We did not observe a change in anti-BDBV GP antibody MFI values at 21 days or 6 months post-vaccination. However, we did observe a remarkable rise in mean BDBV GP reactivity from 1238 to 4845 MFI (p<.0001) between the year 2.5 and year 3.5 visits (Figure 2, top panel).

**Figure 2.**
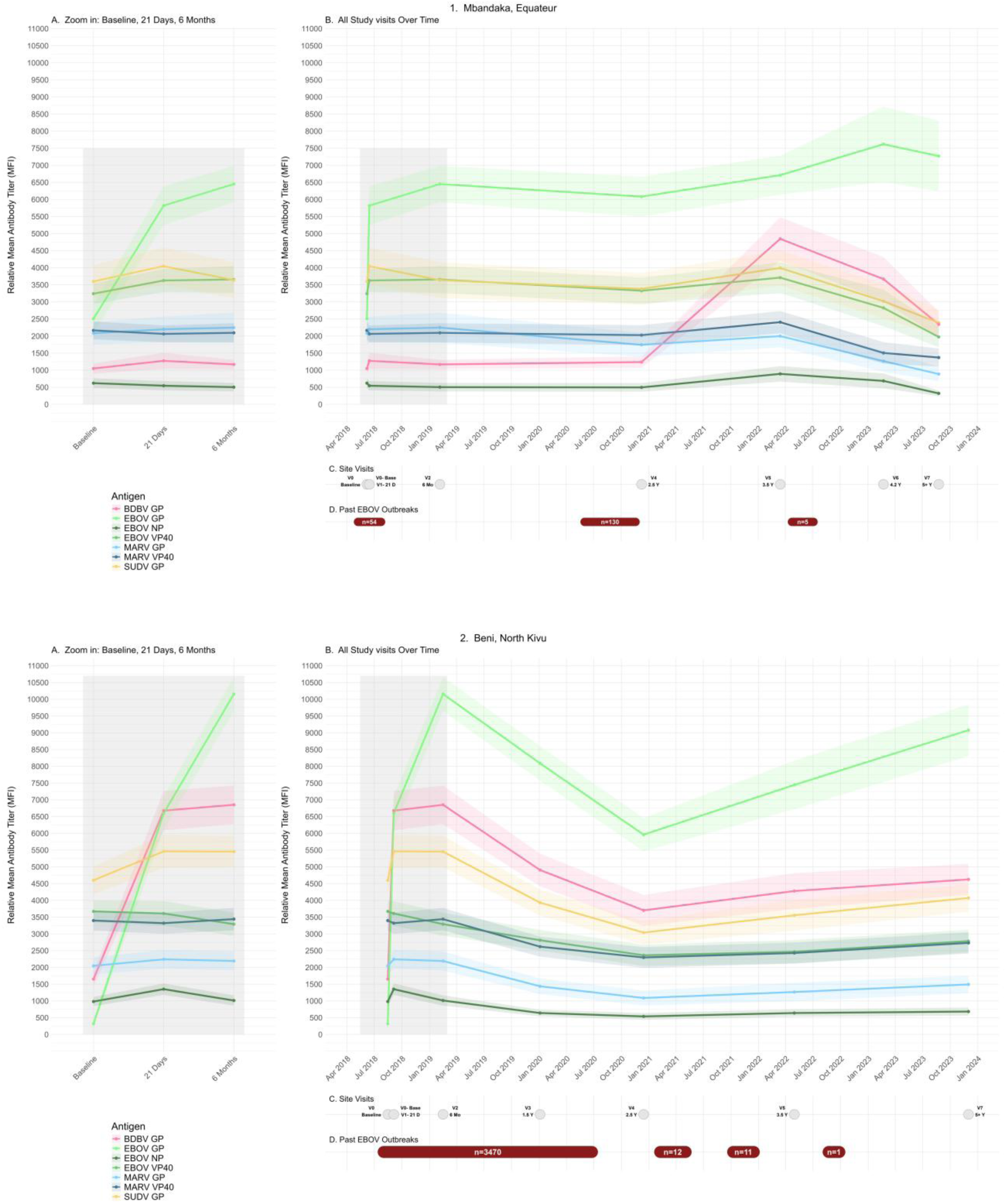
Detected anti-Filovirus Antibody MFI values post rVSVΔG-ZEBOV-GP Vaccination and Outbreak and Vaccine Response Timelines. Anti-filovirus antibody MFI values detected post rVSVΔG-ZEBOV-GP vaccination. Panel 1: Mbandaka, Equateur is plotted above Panel 2: Beni, North Kivu. Sub-panel B shows the mean antibody MFI values and 95% confidence bands to 7 filovirus targets across 7 study visit timepoints plotted over time from. On the left, sub-panel A shows a zoomed-in plot of the section noted in grey-the first three study time points: baseline, 21 days, and 6 months post-vaccination. Below, sub-panel C plots study visit time points (grey boxes), and sub-panel D plots recorded EVD outbreaks (all identified as EBOV) and case counts (red band) in DRC.

Additionally, at the 3.5 year visit, 35.3% (n=116) of participants were seroreactive based on our cut-off criteria (Figure 3). Following this rise we observed a statistically significant waning in anti-BDBV-GP reactivity at the 4.2 year and 5 year visit. At the 3.5 year visit, we observed an uptick in mean MFI values to all filovirus targets, however EBOV NP was the only target other than BDBV GP to have a statistically significant change between 2.5 and 3.5 year visits (Supplemental Figure 1). Among those seroreactive to BDBV GP at 3.5 years who were not seroreactive at baseline (n=109) we observed a mean fold change increase of 14.2 (Supplementary Table 2). At the 3.5 year visit 73.3% (n=241) of participants were reactive to EBOV GP. The mean fold change to EBOV GP from baseline to year 3.5 among those naive at baseline was 4158.2.

**Figure 3.**
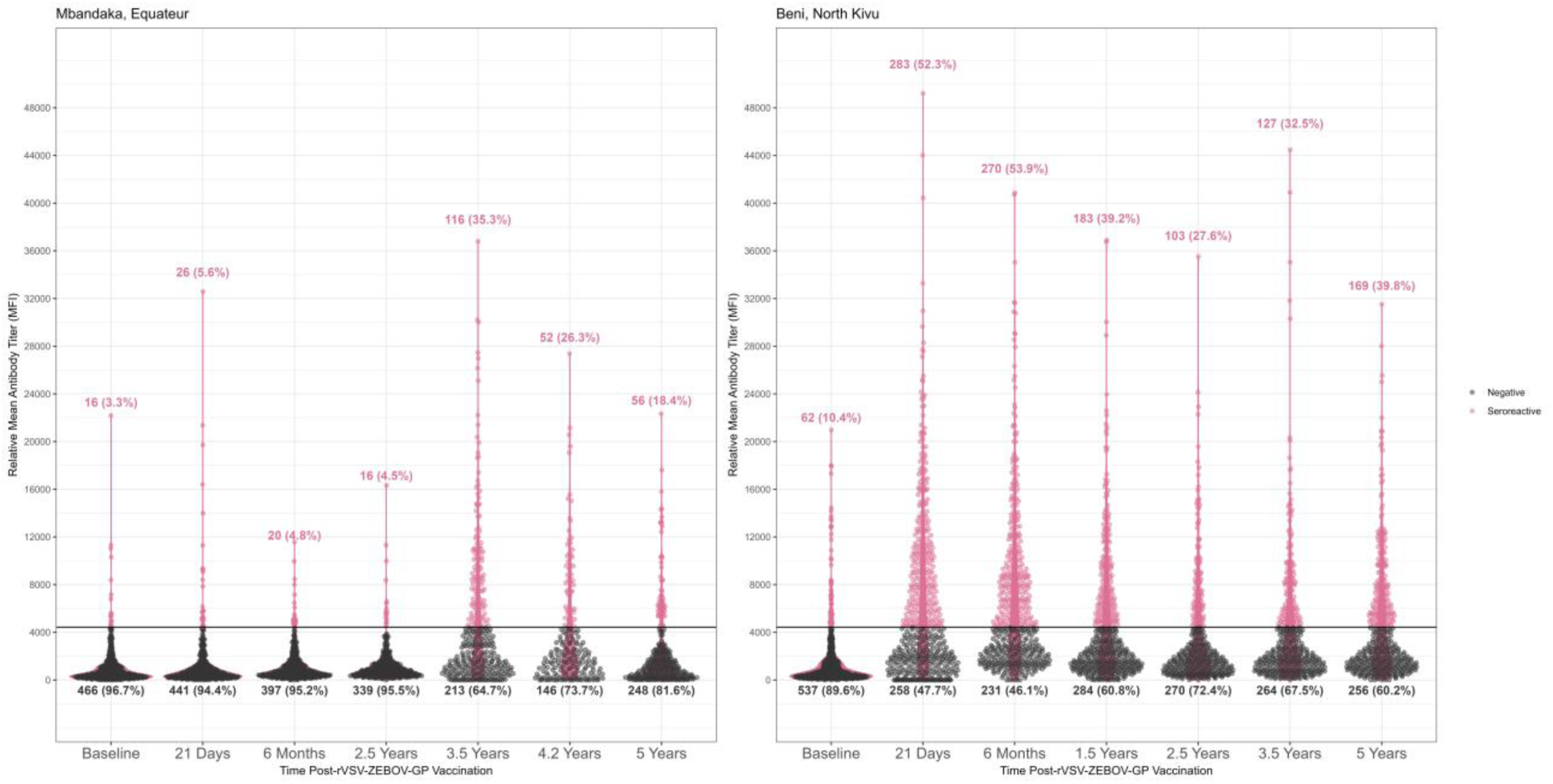
Detected anti-BDBV Antibody MFI values post rVSVΔG-ZEBOV-GP Vaccination. Detected anti-BDBV Antibody MFI values post rVSVΔG-ZEBOV-GP Vaccination. This plot shows the relative mean anti-BDBV GP antibody titer (MFI) among participants in Mbandaka, Equateur (left) and Beni, North Kivu (right) over time. Seroreactive specimens are in pink, sero-negative specimens are in black. A y-intercept line at 4,433 denotes the MFI minimum for seroreactivity. Counts and percentages for seroreactive and seronegative groups are plotted for each timepoint.

In the **Beni cohort,** we observed baseline reactivity to be 10.4% (n=62) to BDBV GP and 3.8% (n=23) to EBOV GP. Interestingly, we observed an immediate rise in anti-BDBV GP antibody MFI values at 21 days and 6 months after vaccination with rVSVΔG-ZEBOV-GP from a mean of 1653 MFI to 6678 (p<.0001). At 21 days and 6 months post-vaccination we observe 52.3% (n=283) and 53.9% (n=270) seroreactivity to BDBV GP, respectively. Among those seronegative to BDBV GP at baseline and seroreactive at 21 days (n=241, 40.2%) and 6 months (n= 227, 37.9%), we recorded a mean fold change of 21.9 and 20.1 compared to baseline, respectively. Mean antibody MFI values decreased for all antigens during the 6 month to 2.5 year post-vaccination period, with only those for EBOV and BDBV GP remaining above the mean MFI values at baseline (Figure 2, bottom panel). The decrease in mean MFI was statistically significant for all filovirus targets between the 6 month to 1.5 year visit. After the 2.5 year study visit we observed a rise in mean MFI in EBOV GP, BDBV GP, and SUDV GP values–between 2.5 years and 5 years this rise was statistically significant (p< .01) (Supplemental Figure 1).

When assessing anti-BDBV GP mean MFI over time for key demographic variables we observed a significant correlation between sex and mean MFI– overall men had higher BDBV GP-reactive mean MFI values compared to women (Supplemental Table 1). We did not observe much variation in mean BDBV GP MFI by age category occupation groups or by vaccination inclusion criteria. However, as several participants met multiple vaccination criteria these categories are not mutually exclusive and thus collinear.

## DISCUSSION

While this investigation and others have demonstrated potent immune responses against EBOV-GP among those vaccinated with rVSVΔG-ZEBOV-GP, it is unknown whether vaccination induces cross-reactive antibodies or confers cross-protection against other *Orthoebolavirus* species, including BDBV.

We detected BDBV GP seroreactivity prior to vaccination, particularly in Beni, where 10.4% of participants were seroreactive at baseline. This DRC-Uganda border region has experienced several EBOV, BDBV, and SUDV outbreaks dating back in the record to 2000. Further investigation is necessary to understand the impact that the endemic nature of these filoviruses may have on underlying lifecourse exposures, background seroreactivity and anti-filovirus antibody MFI values, and most importantly, how background exposures and baseline reactivity may affect vaccine-mediated immune responses.

When considering immunogenicity immediately following vaccination, we identified discordant responses between the Mbandaka and Beni population cohorts. We observed a large increase in anti-BDBV GP MFI values and percent seroreactive at 21 days/6 months post-vaccination in the Beni cohort, significant increases in the MFI values were also observed for EBOV VP40 and SUDV GP. In Mbandaka we did not observe an increase in anti-BDBV GP antibody MFI values at 21 days post-vaccination, or at the next two study time points, which would suggest that the rVSVΔG-ZEBOV-GP vaccine does not elicit strong cross-reactive antibodies to BDBV as compared to EBOV. This finding raises several questions about the nature of these two populations, which are separated geographically by over 1,500km of Congo Basin forest, have differing natural histories of prior and underlying exposures to *Orthoebolaviruses* (known and unknown) and other infections, have differing genetic predispositions and any potential limitations in the laboratory methods that may affect these findings. While anti-BDBV GP MFI values among the Mbandaka cohort remained steady following baseline vaccination, a sharp increase in BDBV GP reactivity was observed during the March 2022 study visit, seroreactivity at this timepoint spiked to 35.3% but then fell to 26.3% and 18.4% at the 4.2 year and 5 year visit. Additionally, analysis should be completed to further look at if there were additional changes for other antigens and if those changes were isolated to specific individuals or more generalized.

While the reasons for these trajectories cannot be determined from the present study, potential explanations include variation in exposure histories prior to and during the study period, environmental factors, undocumented filovirus exposures, differences in immune priming before vaccination, or high non-specific background rates unrelated to filoviruses and immune response post-vaccination. Indeed, several publications have found evidence indicating subclinical or undetected circulation of filoviruses in the DRC^19,24–28^. EVD vaccination in our study regions has been administered in patchwork fashion and comprehensive vaccination records at the ministry-level and individual-level are difficult to verify. Ring vaccination activities took place from 21-May-2018 to 20-Dec-2018 in Mbandaka and from 8-Aug-2018 to 21-Jun-2020 in Beni and multiple booster vaccination activities took place towards the end of our study, but records regarding the timing and location of these activities are sparse. While participants were asked at each study visit participants if they received a EVD vaccine, no public record exists to verify these responses.

This multiplex platform provides efficient evaluation of antibody reactivity across multiple filovirus antigens, yet, serologic assays have many limitations. First, antibody binding measured using a multiplex immunoassay does not establish functional antibody responses such as virus neutralization or Fc-mediated effector functions, and cannot predict a level of protection against infection or disease. Second, the origins of baseline BDBV GP seroreactivity in Beni cannot be determined from these data alone. Third, the timing of seroconversion events occurring between study visits cannot be precisely defined because specimens were collected at discrete intervals and not in response to certain environmental or clinically confirmed incidents. Fourth, there were multiple lots for mastermix for the beads used, however, the same positive control was used across all lots for comparability. Finally, serologic responses were assessed at a single dilution that may not have fallen in the linear range of detectability and therefore may not allow for direct quantitative assessment or functional characterization. Further investigation should include serial dilution of high and low responders to better qualify the upper and lower limits of each assay component in the study setting.

The public health implications of these findings are particularly relevant given the ongoing BDBV outbreak in northeastern DRC and the absence of licensed BDBV-specific vaccines. Although detection of binding antibodies should not be interpreted as evidence of cross-protection, these data expand the limited human and non-human primate evidence base available to evaluate existing Ebola countermeasures against non-EBOV *Orthoebolaviruses* ^29^. Our findings provide longitudinal observations from vaccine recipients residing in regions with recurrent emergence of filoviruses and who qualified for vaccination due to association with a previously detected outbreak. Such data may help inform vaccine recommendations and future studies evaluating the role of licensed EBOV vaccines during outbreaks caused by BDBV or other filoviruses and contribute to ongoing efforts to develop broadly protective filovirus countermeasures. While the functional significance of these responses remains unknown, these findings suggest that some proportion of the population fitting a limited risk profile for receiving vaccines in Northeastern DRC at risk of multiple *Orthoebolaviruses* including EBOV and BDBV may benefit from EBOV vaccination while development efforts towards BDBV-specific vaccines with demonstrated clinical efficacy are ongoing.

## Disclaimer

The views and conclusions contained in this document are those of the authors and should not be interpreted as necessarily representing the official policies, either expressed or implied, of the U.S. Department of Health and Human Services or of the institutions and companies affiliated with the authors, nor does mention of trade names, commercial products, or organizations imply endorsement by the U.S. Government. This project has been funded in whole or in part with Federal funds from the Food and Drug Administration (Grant No. 75F40119C10128), the Gates Foundation (OPP1195609), and U.S. Defense Advanced Projects Agency (DARPA, N66001-09-C-2082 and HR0011-17-2-0069).

## Data Availability

Data produced in the present study are available upon reasonable request to the authors.

---

**Supplemental Figure 1:**
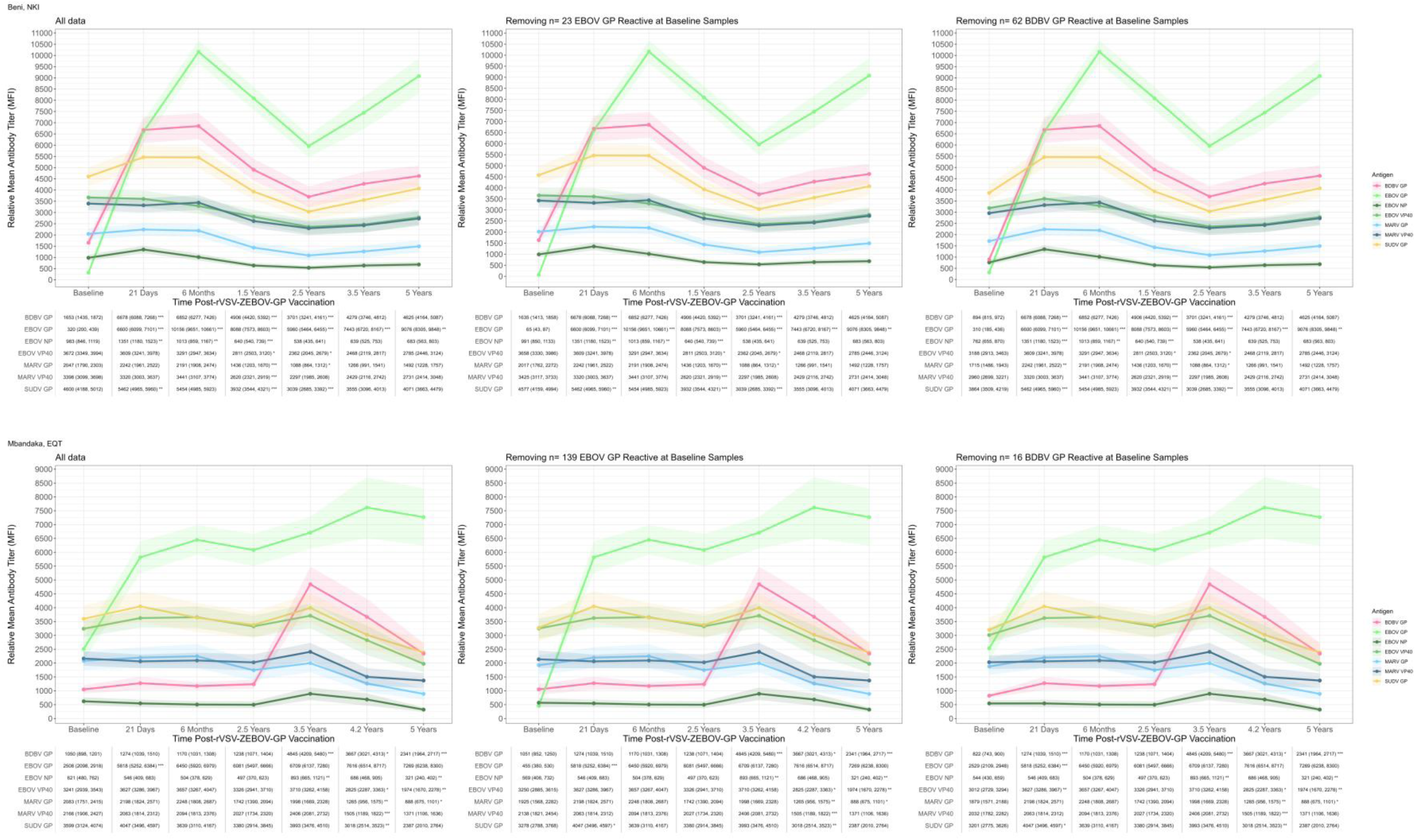
Reactivity to Filovirus Antigens over Five-Year follow up, censoring those individuals reactive at baseline to EBOV GP or BDBV GP. Anti-filovirus antibody MFI values detected post rVSVΔG-ZEBOV-GP vaccination. Mbandaka, Equateur is plotted above Beni, North Kivu. Below each plot is a table of the mean MFI values and the 95% confidence interval for each filovirus antigen. Asterisks denote the t-test p-value comparing the mean MFI of that visit to the mean MFI of the previous visit, * = <0.05, ** = <0.01, *** = <.0001, Column 1 labeled “all data” plots all observations and is the same plot represented in Figure 1. The middle column removes individuals who were EBOV GP reactive at baseline. The right most column plots the mean MFI values removing those who were BDBV reactive at baseline

**Supplemental Figure 2:**
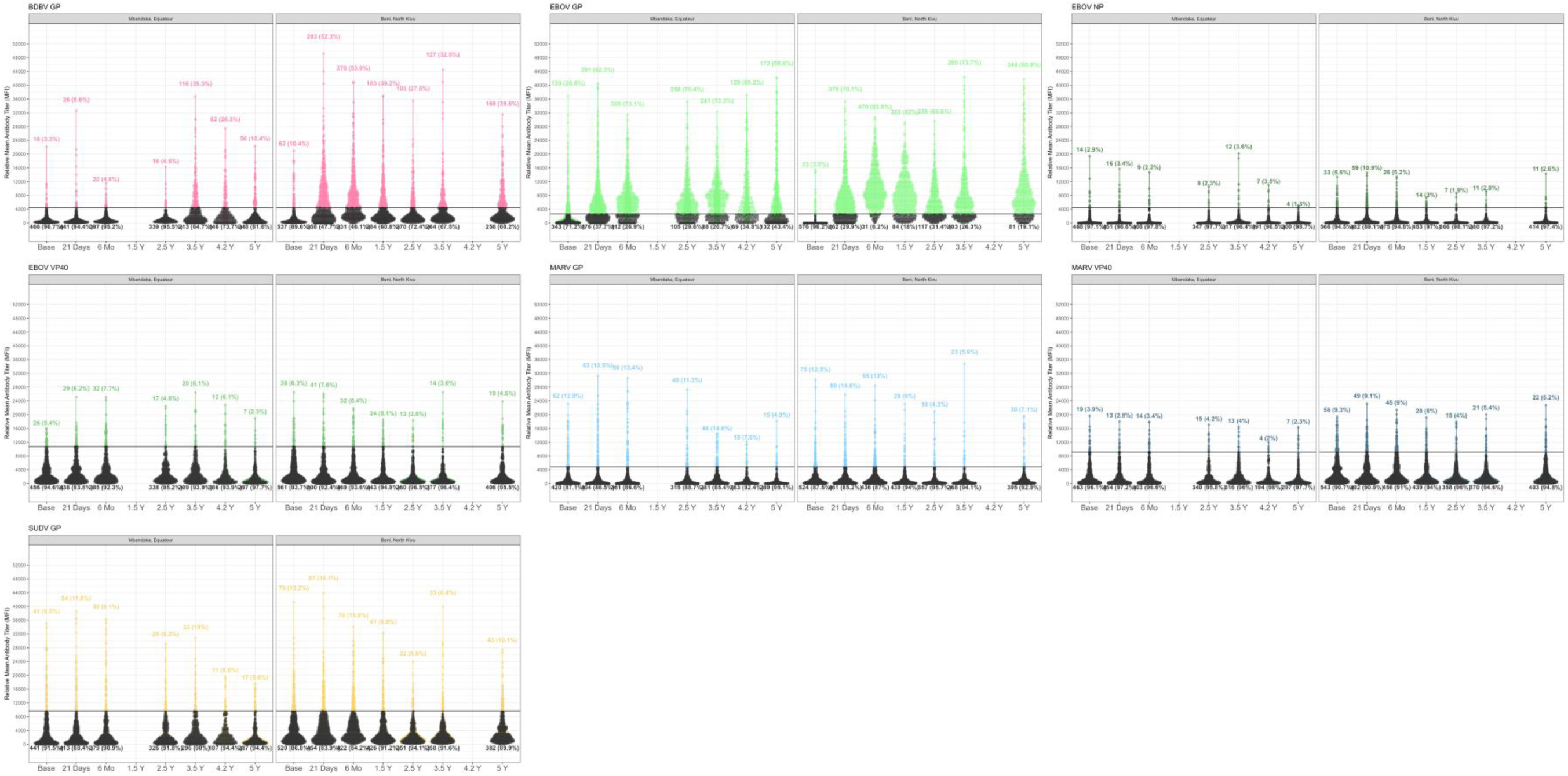
Detected Antibody MFI values post rVSVΔG-ZEBOV-GP Vaccination for seven filovirus antigens. Horizontal line is the calculated cut-off as described in methods.

**Supplementary Table 1.**
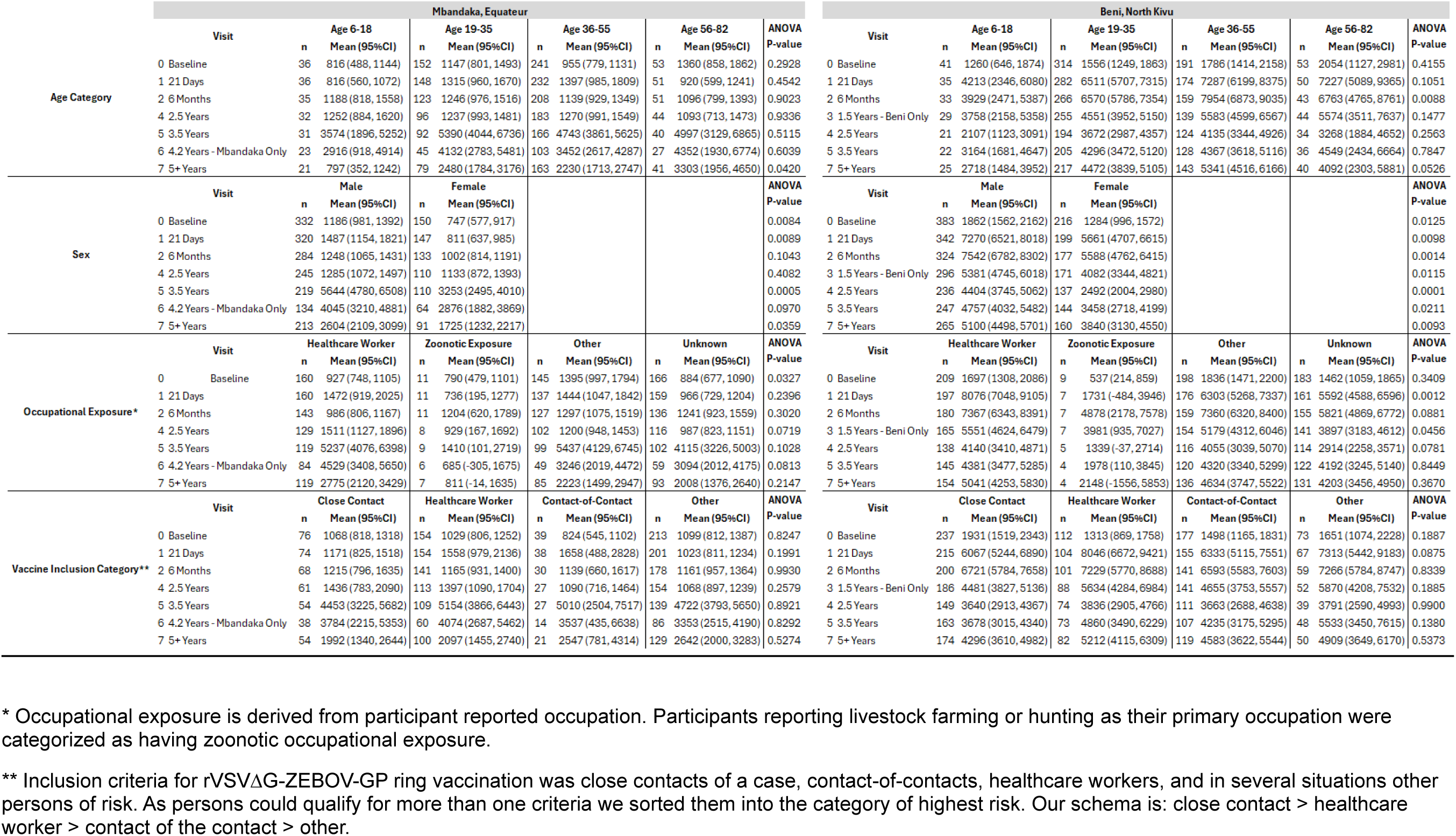
Mean BDBV GP MFI across visits by Demographic Characteristic.

**Supplemental Table 2.**
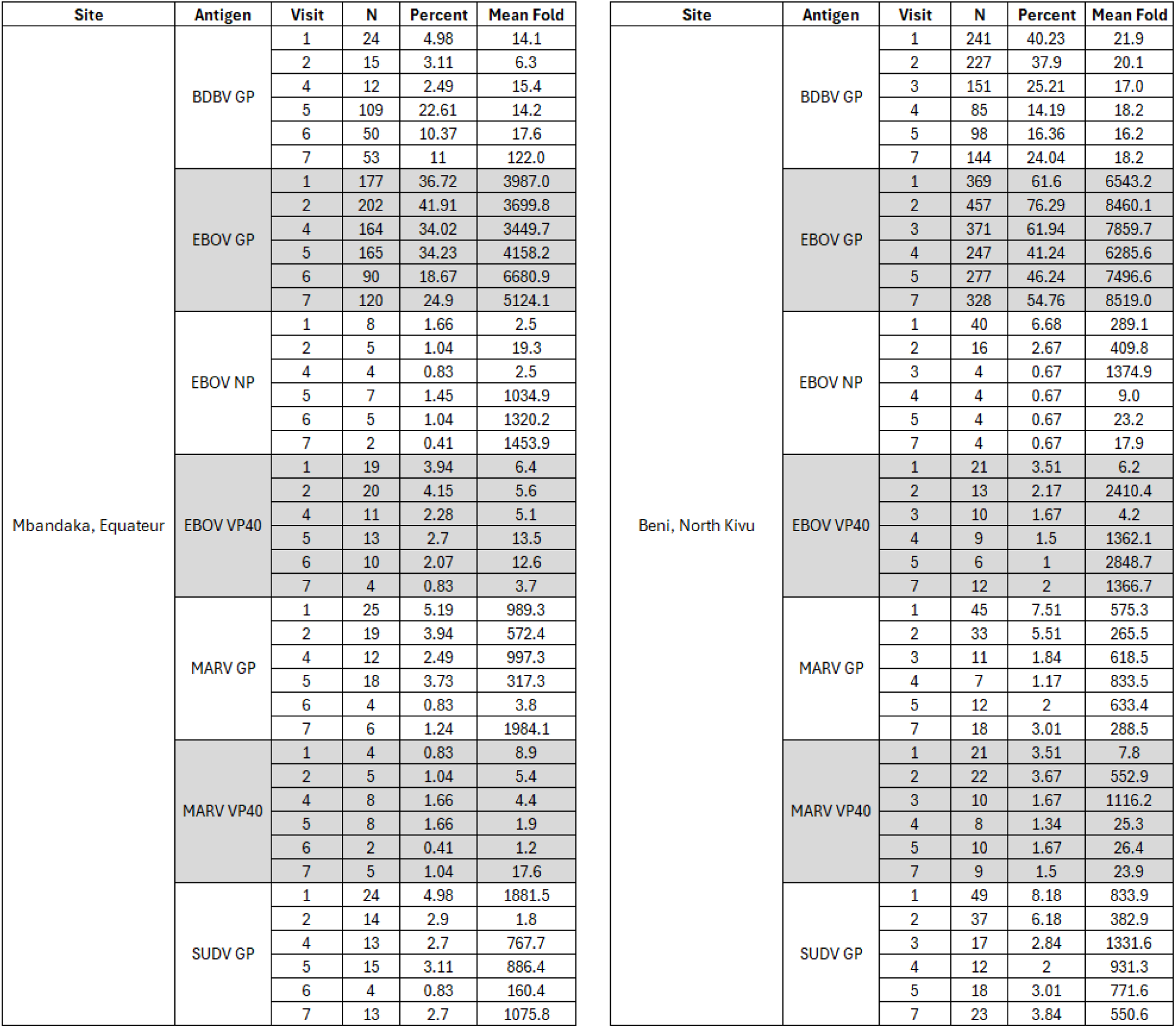
Fold change. Fold change for each individual is calculated as the change since baseline (visit = 0). Mean fold changes were calculated by grouping only individuals who were seroreactive at each visit and excluding those seroreactive at baseline. N is the count of individuals meeting this criteria for each visit and antigen, Percent is out of the total persons enrolled in each cohort, N= 482 in Mbandaka, Equateur and N= 599 in Beni, North Kivu.

## References

1. WHO. Epidemic of Ebola Disease caused by Bundibugyo virus in the Democratic Republic of the Congo and Uganda determined a public health emergency of international concern. 2026; https://www.who.int/news/item/17-05-2026-epidemic-of-ebola-disease-in-the-democratic-republic-of-the-congo-and-uganda-determined-a-public-health-emergency-of-international-concern. Accessed June 22, 2026.

2. Phelan AL, Nuzzo JB, Gostin LO. The PHEIC for Ebola disease caused by Bundibugyo virus: an inflection point for solidarity and health equity. The Lancet. 2026;407(10545):2264–2267.

3. Henao-Restrepo AM, Camacho A, Longini IM, et al. Efficacy and effectiveness of an rVSV-vectored vaccine in preventing Ebola virus disease: final results from the Guinea ring vaccination, open-label, cluster-randomised trial (Ebola Ça Suffit!). Lancet. 2017;389(10068):505–518.

4. Henao-Restrepo AM, Longini IM, Egger M, et al. Efficacy and effectiveness of an rVSV-vectored vaccine expressing Ebola surface glycoprotein: interim results from the Guinea ring vaccination cluster-randomised trial. Lancet. 2015;386(9996):857–866.

5. Rupani N, Ngole ME, Lee JA, et al. Effect of Recombinant Vesicular Stomatitis Virus-Zaire Ebola Virus Vaccination on Ebola Virus Disease Illness and Death, Democratic Republic of the Congo. Emerg Infect Dis. 2022;28(6):1180–1188.

6. Coulborn RM, Bastard M, Peyraud N, et al. Case fatality risk among individuals vaccinated with rVSVΔG-ZEBOV-GP: a retrospective cohort analysis of patients with confirmed Ebola virus disease in the Democratic Republic of the Congo. Lancet Infect Dis. 2024;24(6):602–610.

7. Kasereka MC, Ericson AD, Conroy AL, Tumba L, Mwesha OD, Hawkes MT. Prior vaccination with recombinant Vesicular Stomatitis Virus - Zaire Ebolavirus vaccine is associated with improved survival among patients with Ebolavirus infection. Vaccine. 2020;38(14):3003–3007.

8. Meakin S, Nsio J, Camacho A, et al. Effectiveness of rVSV-ZEBOV vaccination during the 2018-20 Ebola virus disease epidemic in the Democratic Republic of the Congo: a retrospective test-negative study. Lancet Infect Dis. 2024;24(12):1357–1365.

9. WHO. *Preliminary results on the efficacy of rVSV-ZEBOV-GP Ebola vaccine using the ring vaccination strategy in the control of an Ebola outbreak in the Democratic Republic of the Congo: an example of integration of research into epidemic response*. 2019.

10. North Kivu/Ituri, Democratic Republic of the Congo, August 2018 - June 2020 [press release]. 2020.

11. Mdluli T, Wollen-Roberts S, Merbah M, et al. Ebola virus vaccination elicits Ebola virus–specific immune responses without substantial cross-reactivity to other filoviruses. Science Translational Medicine. 2025;17(792):eadq2496.

12. Lhomme E, Wiedemann A, Ayouba A, et al. Cross-reactive Bundibugyo antibody responses after licensed Ebola vaccines. medRxiv. 2026.

13. Ajelli M, Muyembe J-J, Touré A, et al. Vaccination strategies for Ebola in the democratic republic of Congo: the WHO-Ebola modeling collaboration. International Journal of Infectious Diseases. 2025;153:107779.

14. Program WHE. Ebola Virus Disease, Democratic Republic of the Congo. 2018.

15. Ebola virus disease – Democratic Republic of the Congo [press release]. 2018.

16. Hoff NA, Bratcher A, Kelly JD, et al. Immunogenicity of rVSVΔG-ZEBOV-GP Ebola vaccination in exposed and potentially exposed persons in the Democratic Republic of the Congo. Proc Natl Acad Sci U S A. 2022;119(6).

17. Control CfD. History of Ebola Outbreaks. https://www.cdc.gov/ebola/outbreaks/index.html, 2026.

18. Becton DaC. BD Vacutainer® serum tubes. In. Franklin Lakes, NJ2026.

19. Merritt S, Mukadi PK, Kompany JP, et al. Detection of Marburg Virus Antibodies 25 Years After Outbreak in Watsa, Democratic Republic of the Congo. The Journal of Infectious Diseases. 2026.

20. Lehrer AT, Wong T-AS, Lieberman MM, et al. Recombinant proteins of Zaire ebolavirus induce potent humoral and cellular immune responses and protect against live virus infection in mice. Vaccine. 2018;36(22):3090–3100.

21. Haun BK, Kamara V, Dweh AS, et al. Serological evidence of Ebola virus exposure in dogs from affected communities in Liberia: A preliminary report. PLOS Neglected Tropical Diseases. 2019;13(7):e0007614.

22. Merritt S, Halbrook M, Kompany JP, et al. Comparison of EBOV GP IgG antibody reactivity: Results from two immunoassays in the Democratic Republic of the Congo. Journal of Virological Methods. 2025;336:115154.

23. Merritt S, Hoff NA, Mukadi PK, et al. Sustained Specific EBOV GP Immunogenicity Five-Years Post-Vaccination: Longitudinal Results from North Kivu and Equateur, Democratic Republic of the Congo. medRxiv. 2026:2026.2005.2019.26353493.

24. Semancik CS, Cooper CL, Postler TS, et al. Prevalence of human filovirus infections in sub-Saharan Africa: A systematic review and meta-analysis protocol. Syst Rev. 2024;13(1):218.

25. Bower H, Glynn JR. A systematic review and meta-analysis of seroprevalence surveys of ebolavirus infection. Scientific Data. 2017;4(1):160133.

26. Bratcher A, Hoff NA, Doshi RH, et al. Zoonotic risk factors associated with seroprevalence of Ebola virus GP antibodies in the absence of diagnosed Ebola virus disease in the Democratic Republic of Congo. PLOS Neglected Tropical Diseases. 2021;15(8):e0009566.

27. Crozier I. Mapping a Filoviral Serologic Footprint in the Democratic Republic of the Congo: Who Goes There? J Infect Dis. 2018;217(4):513–515.

28. Mulangu S, Alfonso VH, Hoff NA, et al. Serologic Evidence of Ebolavirus Infection in a Population With No History of Outbreaks in the Democratic Republic of the Congo. J Infect Dis. 2018;217(4):529–537.

29. Falzarano D, Feldmann F, Grolla A, et al. Single immunization with a monovalent vesicular stomatitis virus-based vaccine protects nonhuman primates against heterologous challenge with Bundibugyo ebolavirus. J Infect Dis. 2011;204 Suppl 3(Suppl 3):S1082–1089.

